# Impact of exposure to per- and polyfluoroalkyl substances on fecal microbiota composition in mother-infant dyads

**DOI:** 10.1101/2022.12.14.22283359

**Authors:** Santosh Lamichhane, Taina Härkönen, Tommi Vatanen, Tuulia Hyötyläinen, Mikael Knip, Matej Orešič

## Abstract

Current evidence suggests that chemical exposure alters gut microbiota composition, with higher exposure to environmental chemicals being associated with reduced microbiome diversity. However, not much is known about the impact of per- and polyfluoroalkyl substances (PFAS) on gut bacteria. Here we set out to identify the gut bacterial species that associate with chemical exposure before (maternal) and after (maternal, infant) birth in a mother-infant series. Paired blood and stool samples were collected from mother-infant dyads (n = 30) in a longitudinal setting. PFAS were quantified in maternal blood to examine their associations with the microbial compositions (determined by shotgun metagenomic sequencing) in mothers and infants. High maternal exposure to PFAS was persistently associated with increased abundance of *Methanobrevibacter smithii* in maternal stool. Among individual PFAS compounds, PFOS and PFHpS showed the strongest connection with *M. smithii*. However, maternal PFAS exposure associated only weakly with the infant microbiome. Our findings suggest that PFAS exposure contributes to the modulation of the adult gut microbiome composition.

## Introduction

Human gut harbors a complex and highly dynamic microbial ecosystem that is conspicuously variable within and between individuals (Gilbertet al. 2018; Turnbaughet al. 2007). Gut microbes are involved in a multitude of host physiological functions including immune response, host metabolism, and nervous system development (Gilbertet al. 2018). Changes in the composition of gut microbes have been associated with many human diseases, from diabetes to mental health (Ruuskanenet al. 2022; Valles-Colomeret al. 2019). Host genotype and the non-genetic factors such as diet, lifestyle, and xenobiotic exposure are crucial in shaping the human gut microbiome composition throughout life (Rothschildet al. 2018), which may, however, results in various adverse health outcomes (Smitset al. 2017).

Humans are exposed to many environmental chemicals. Out of more than 140,000 industrially produced chemicals, humans are exposed to about 5000, out of which over 400 chemicals or their metabolites are detectable in human biological samples (CDC 2022; Vermeulenet al. 2020). One specific class of persistent environmental chemicals that humans are widely exposed to include per- and polyfluoroalkyl substances (PFAS), with their biological half-lives ranging from three to five years (Liet al. 2018; Liewet al. 2018). Human exposure to PFAS predominantly takes place through ingestion of contaminated food and drinking water (Sunderlandet al. 2019). Fetuses are already exposed to PFAS *in utero via* placental transfer and in infants the cumulative exposure is transmitted *via* maternal breastmilk (Croeset al. 2012; Lamichhaneet al. 2021).

The human gut microbiome contributes to the metabolism of ingested xenobiotic compounds (Koppelet al. 2017; Sunderlandet al. 2019). Thus, exposure to PFAS in early life may affect the maturation of the gut microbiome and the host immune system. Current evidence suggests that high exposure to PFAS during early life can lead to adverse health outcomes in adulthood (Sunderlandet al. 2019). High PFAS exposure has been shown to reduce microbiome diversity in infants (Iszattet al. 2019). While several studies have demonstrated an association between exposure to a series of various environmental chemicals (bisphenols, phthalates, persistent organic pollutants, heavy metals, and pesticides) and gut microbiome, a significant knowledge gap persists regarding how PFAS influence the overall viability of gut microbes (Chiuet al. 2020). Importantly, we do not yet understand the potential of PFAS to alter the human gut microbiome at the individual species level. Here, we aimed to identify the gut microbial species that associate with PFAS exposure prenatally and during early life in a mother-child cohort.

## Results

### Metabolomics analyses of the mother-infant cohort

Thirty mother-child dyads were included in the study (Fig.1, Table S1). Serum concentrations of 20 PFAS compounds were quantified in maternal serum samples drawn during pregnancy. Serum metabolites were quantified in serum samples collected at 3 months after the delivery from the mothers and the offspring. We analyzed metabolites from across a wide range of chemical classes, including, bile acids, amino acids, free fatty acids, and other chemical pollutants including benzyl paraben and furoic acid (CMPFPA). Relative abundance patterns of microbial species were characterized by shotgun metagenomics sequencing in longitudinal mother and child stool samples: Infant –birth (meconium) and 3 months of age; Mother – gestational (G, during pregnancy), delivery (D), and three months post-delivery (T).

**Figure 1.**
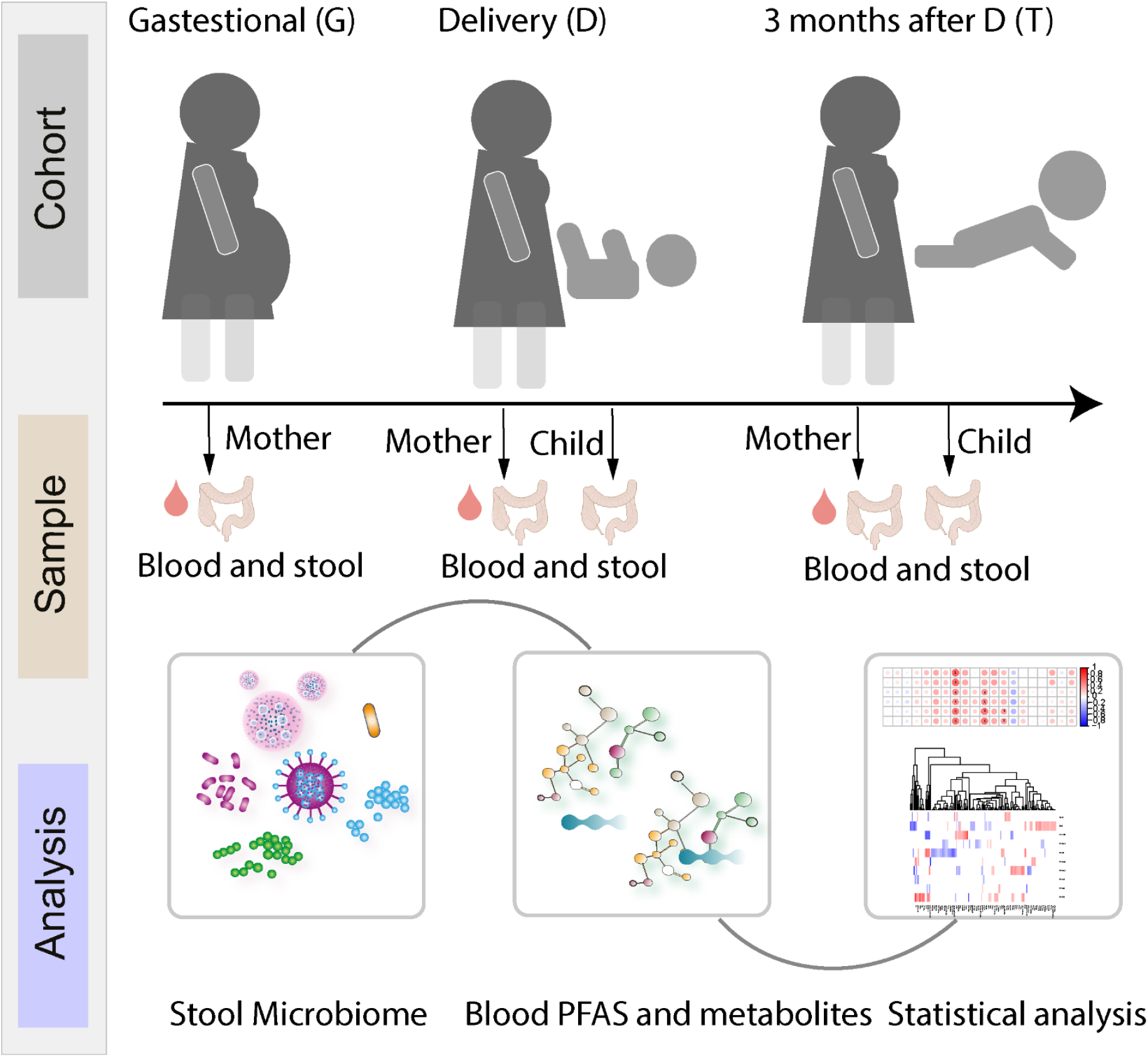
An overview of the study setting. Longitudinal mother and child stool sample from five time points: Two offspring samples at birth (meconium), and at 3 months of age, together with three maternal stool samples: gestational (G, during pregnancy at the beginning of the third trimester), delivery (D), and 3 months after the delivery (T). Paired serum and stool samples from mothers and offspring were collected at the infant age of 3 month. Selected characteristics of the study subjects are shown in Table S1.

### Impact of PFAS exposure on the gut microbiome

PFAS exposure associated with the gut microbiome profile (Fig. 2). To understand the impact of PFAS on the microbial communities in the mothers and in the offspring, we classified the microbiome profile into two groups (high vs. low exposure) based on total maternal PFAS exposure levels (quartiles), with the data being analyzed separately for mother and child at any individual time point. We performed linear discriminant effect size (LEfSe) analysis (Segataet al. 2011) to determine the species-level differences between the highest (Q3+Q4) and lowest (Q1+Q2) PFAS exposure quartiles.

**Figure 2.**
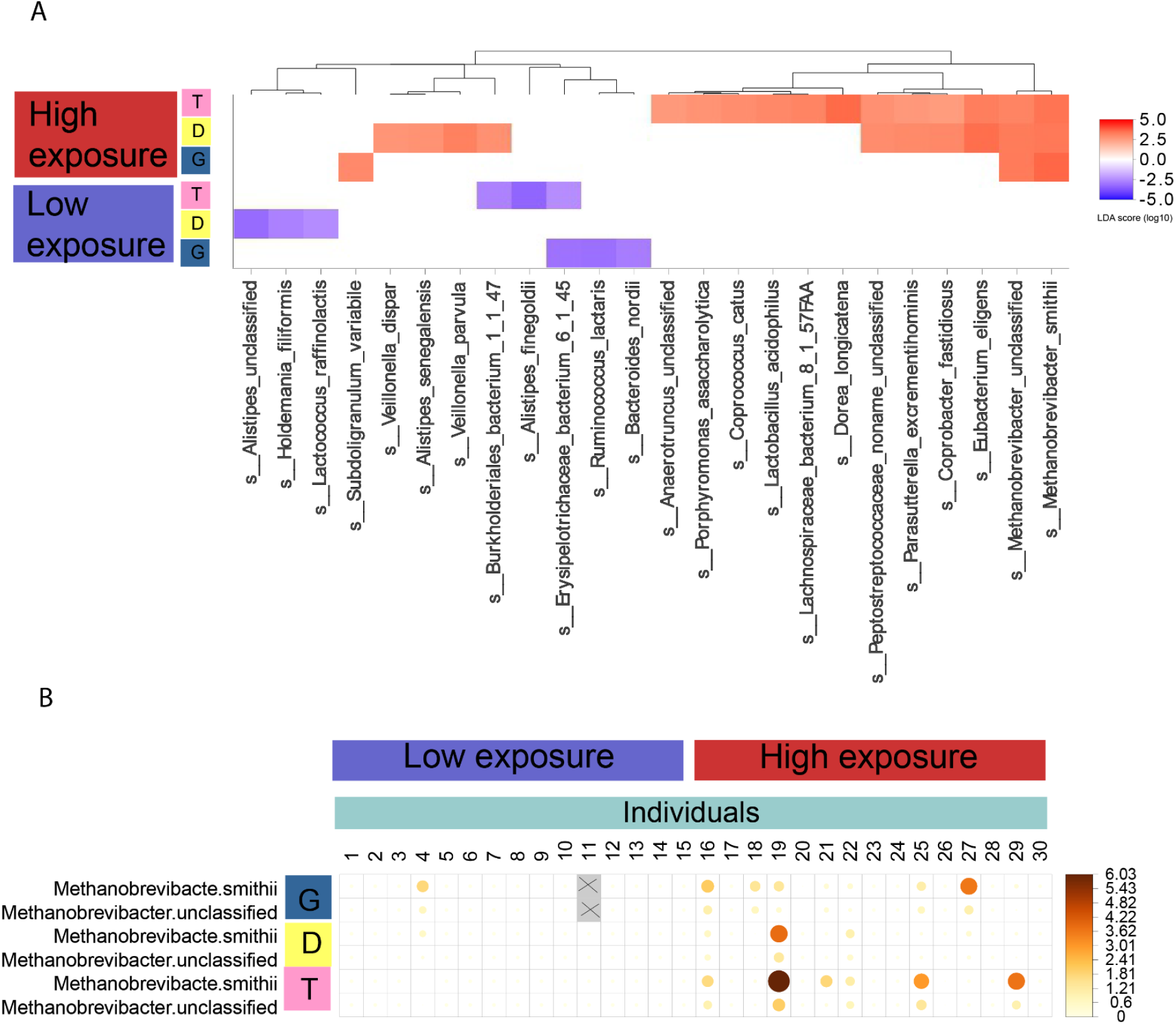
Comparison of microbiome abundances between the exposure groups. a) Heat map showing the log-transformed linear discriminant analysis (LDA) scores computed with linear discriminant analysis effect size (LEfSe) for significantly different bacterial taxa between high vs. low exposure groups at the different sampling time points. The LDA score indicates the effect size and ranking of each microbial taxon. Statistical significance was evaluated using the Wilcoxon rank-sum tests in LEfSe. b) Heat map showing relative abundances of *Methanobrevibacter* species were persistently higher in the highly exposed stool sample in the longitudinal settings. Here, X, shaded light grey represents missing sample collected during pregnancy at the beginning of the third trimester.

We found that during pregnancy, a total of six microbial species differed between higher and lower quartiles of maternal exposure to PFAS (Fig. 2). *Methanobrevibacter smithii*, additional unclassified *Methanobrevibacter* sequences, and *Subdoligranulum variabile* were more abundant in highly exposed group, while *Erysipelotrichaceae bacterium 6145, Bacteroides nordii* and *Ruminococcus lactaris* were more abundant in the low exposure group. Comparing high *vs*. low exposure at the time of delivery revealed elevated levels of 10 microbial species (*Burkholderiales bacterium 1147, Veillonella parvula, Alistipes senegalensis, Coprobacter fastidiosus, Parasutterella excrementihominis, Eubacterium eligens, Peptostreptococcaceae noname unclassified, Veillonella dispar, Methanobrevibacter smithii, Methanobrevibacter unclassified*) in the highly exposed mothers (Fig. 2). Similarly, we compared the maternal gut microbiome difference between the exposures groups in the samples collected 3 months after the delivery. A total of 15 microbial species were different between the high and low exposure groups. There was no persistent trend with respect to microbial differences between the groups (Fig. 2A), with the exception for *Methanobrevibacter* (Fig. 2), which was persistently higher in high exposure group as compared to low exposure.

Infants are highly exposed to PFAS due to cumulative exposure *vi*a breastfeeding during early life. To study the potential impact of PFAS on microbial communities in infants, we compared the maternal PFAS exposure quartiles with the infants’ gut microbiome profiles. We found that maternal PFAS showed only modest association with stool microbiome in the infants. The linear discriminant effect size analysis revealed a total of three microbial species (*Subdoligranulum unclassified, Bifidobacterium pseudocatenulatum*, and *Propionibacterium avidum*), which were altered between birth and at 3 months of age.

Next, we sought to determine associations of individual PFAS on those gut microbes that had different abundances between the high and low exposure maternal groups (Fig. 2). By using ranked based correlation analysis, we tested for associations between 20 measured PFAS species with those microbial species that were different between the high and low exposure group in two or more maternal sampling time points (*i*.*e*., G, D, or T). PFOS and PFHpS were found persistently associated with *Methanobrevibacter* species (Fig. 3). PFECHS showed positive association with *Methanobrevibacter* in the sample collected 3 months post-delivery. In addition, we found that *Erysipelotrichaceae bacterium 6145* was inversely associated with PFOS and PFUnDA during pregnancy, while it remained positively associated with PFPeS at the time of delivery.

**Figure 3.**
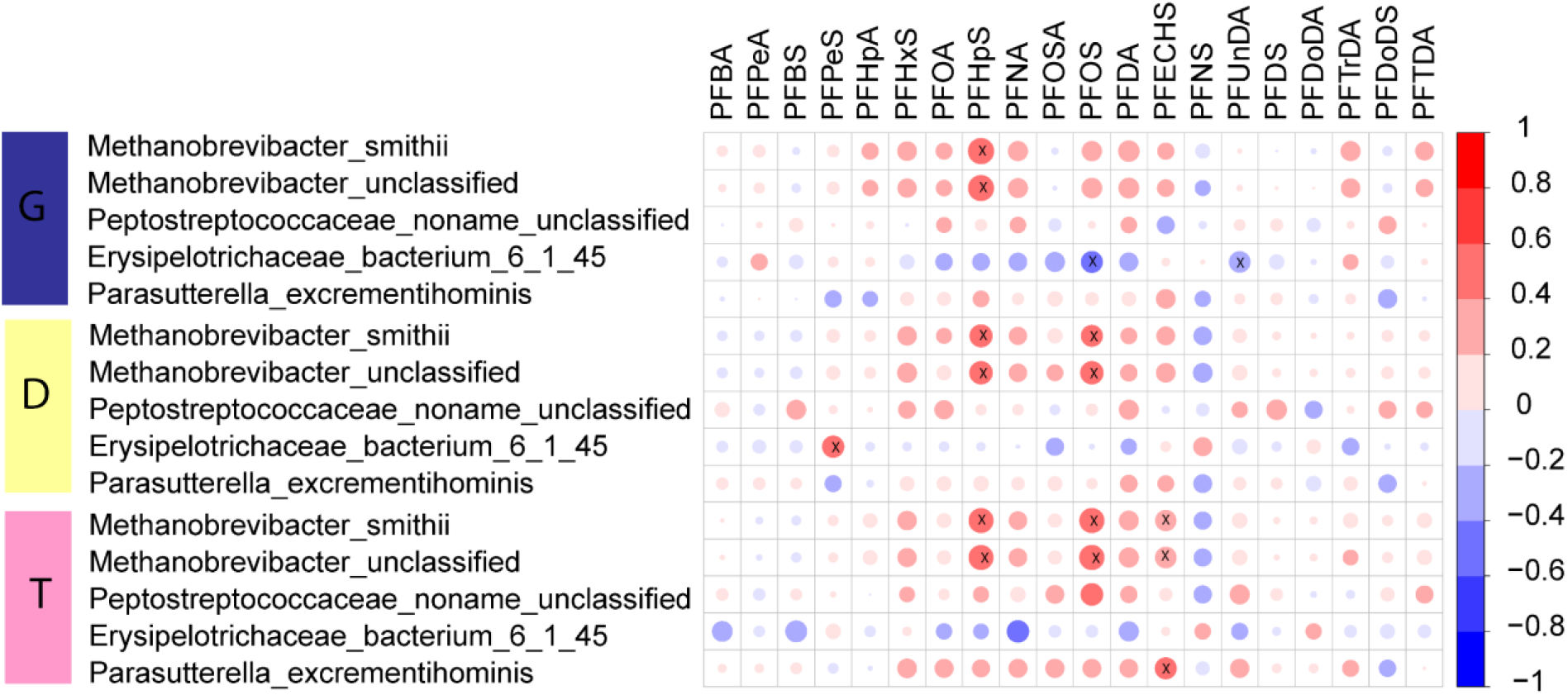
Associations between the microbiome and PFAS in the mothers. Spearman rank correlation between the individual PFAS species (n = 20) with those microbial species that appeared to be affected in a minimum of two longitudinal sampling time points (i.e. G, D, and T) in the mothers.

### Metabolomic profiles in mothers were associated with microbiome profile

We also examined whether the abundance of altered PFAS-associated bacterial species correlated with the levels of circulating metabolites. We found that *Methanobrevibacter* species were inversely associated with glycine-conjugated BAs, particularly with GDCA, GUDCA and GUDCAS (Fig.4). *Alistipes putredinis* was inversely associated with docosahexanoic acid (C22:6) and one BA (isoUDCA or isoHDCA or MCDCA). In the maternal serum at 3 months post-delivery, we also observed positive associations between *Parasutterella excrementihominis* and microbial-derived metabolite indole-3-acetic acid and two BAs (GUDCA and CA). In addition, Erysipelotrichaceae bacterium 6145 showed a significant, positive association with 12-ketolithocholic acid, indicating that *Erysipelotrichaceae* and *Parasutterella* may actively participate in human BA metabolism and homeostasis (Fig.4).

**Figure 4.**
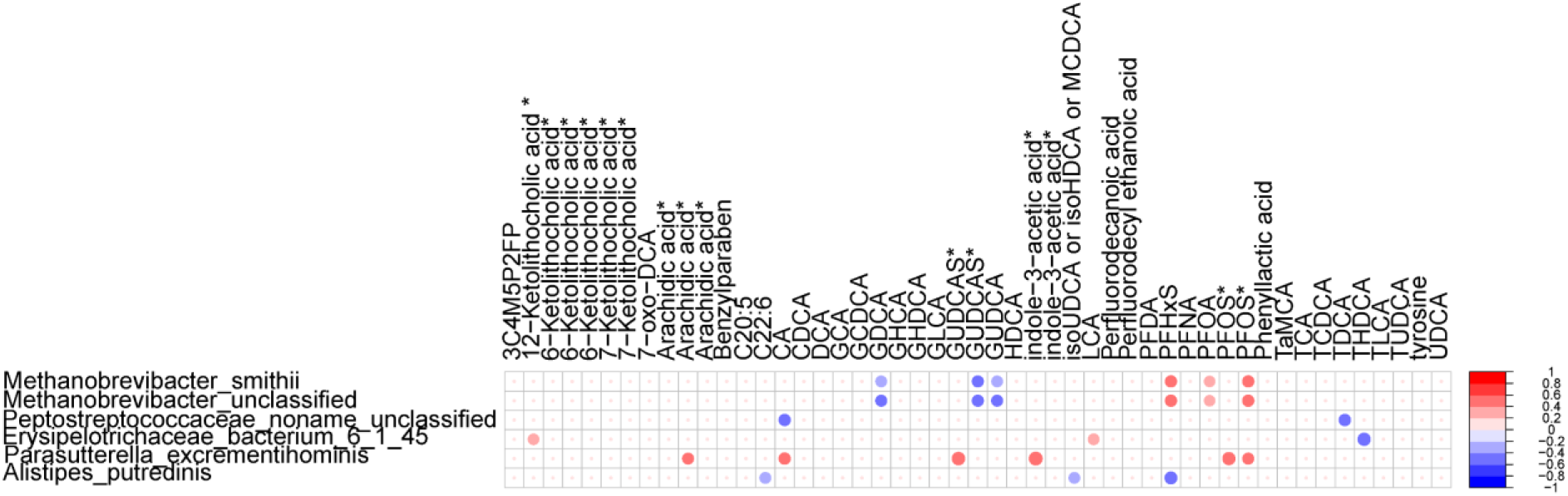
Associations between microbiome and circulatory metabolite in the mothers. Spearman rank correlation between the polar metabolites with those microbial species that eared to be affected in a minimum of two longitudinal sampling time points (i.e. G, D, and T) in the mothers. * indicate the isomers.

## Discussion

In this mother-offspring study, we found that the maternal gut microbiome composition was altered in mothers highly exposed to PFAS, with relative abundances of *Methanobrevibacter* species being persistently higher as compared to unexposed mothers. We were also able to demonstrate that the microbially-derived metabolite indole-3-acetic acid and the conjugated BA (such as GUDCA) were associated with an altered maternal microbiome 3 months after delivery.

Our observations suggest that the diversity and/or abundance of the methanogenic archaea may vary with PFAS exposure, which is in agreement with recent studies reporting that persistent organic exposure induce changes in human microbiome diversity (Iszattet al. 2019; Naspoliniet al. 2022). High maternal exposure to PFAS was previously found associated with a decrease of *Faecalibacterium* and increase of *Clostridium, Staphylococcus*, and *Bifidobacterium* bacterial species in infant stool samples (Naspoliniet al. 2022). In previous studies PFAS exposure was also associated with disturbed stool microbiome diversity in infants, specifically in the bacterial domain (Iszattet al. 2019; Naspoliniet al. 2022).

Here, we focused on mother-child pairs, and the metagenomics sequencing approach allowed us to investigate microbiome samples at subspecies resolution (Yassouret al. 2018). We found that PFAS exposure had profound impact on the *Methanobrevibacter* species in maternal sample but not in the infant microbiome, which could be due to the absence of *Methanobrevibacter* colonization/succession in the infant gut. Interestingly, we identified two specific PFAS, PFOS and PFHpS, which were strongly associated with the *Methanobrevibacter*, a dominant methanogenic archaea in the adult human gut. These methanogens are functionally important due to their ability to consume molecular hydrogen and contribute to host energy harvest (Hansenet al. 2011). Importantly, *Methanobrevibacter* strains have been reported to be proinflammatory pathobionts associated with multiple sclerosis (Jangiet al. 2016), childhood obesity (Mbakwaet al. 2015) and urinary tract infection (Grineet al. 2019). Meanwhile, *Methanobrevibacter* were also found to be enriched in Sardinian centenarians (Wuet al. 2019).

Gut microbiome has a profound influence on the human metabolome (Dekkerset al. 2022). We found that microbial species (archaea) which were affected by PFAS exposure, were inversely associated with glycine-conjugated BAs, in particular with GDCA, GUDCA and GUDCAS. The BA pool in humans is also regulated by the gut microbes, secondary BAs in particular (Ridlonet al. 2014). Functionally, PFAS have been suggested to impact the intestinal re-absorption of BAs (Beggset al. 2016; Chiang 2017; Zhaoet al. 2015). Previous studies indicated that BA and methanogens in the human colon are interlinked (Florin and Jabbar 1994). Recently, gut-associated archaea M. *smithii* have also been found to have bile salt hydrolases (BSHs) activity, which is responsible for conjugation of amino acids to BAs (Joneset al. 2008).

However, our understanding about the role of the methanogenic archaea on BA metabolism is still limited. Therefore, we hypothesize that high PFAS exposure impact the composition and diversity of the gut microbiome, which, in turn, may also affect the BA metabolism or vice versa. Our data do not reveal the cause and effect relationships. However the observed strong association between the altered microbe and conjugated BAs implies that archaea may have a role in mediating the interplay of BA metabolism, re-absorption and the impact of PFAS exposure, which clearly demands further investigation. We also acknowledge that sample size in our study, despite longitudinal setting, may have been too small for gaining a comprehensive view of the impact of chemical exposures on the gut microbiome in a mother-infant dyad setting. Nevertheless, our findings do suggest that PFAS exposure contributes to the modulation of the gut microbiome composition.

## Methods

### Study population

The study cohort of 30 healthy pregnant women were recruited from January 28, 2013 to February 26, 2015 (Yassouret al. 2018). Families were contacted at the fetal ultrasonography visit, which is arranged for all pregnant women in Finland around gestational week 20. Written informed consent was signed by the parents at the beginning of the third trimester to analyze HLA genotype of the offspring. Patient consent was overseen by the Ethical Committee of the Joint Municipal Authority of the Pirkanmaa Hospital District. Inclusion criteria for the study were informed consent signed by the parents and an eligible HLA genotype of the newborn conferring increased risk for type 1 diabetes (T1D), as this cohort is a subset of another larger T1D-centered cohort. Eligible genotypes, determined as previously described (Ilonen et al., 2016) were the high-risk genotype combining (DR3)-DQA1*05-DQB1*02 and DRB1*0401/2/4/5-DQA1*03-DQB1*03:02 haplotypes or the moderate-risk genotypes defined as homozygosity for either one, DRB1*04:01/2/5-DQA1 *03-DQB1 *03:02 with a neutral haplotype, and the (DR3)-DQA1*05-DQB1*02/DRB1*09-DQA1*03-DQB1*03:03 genotype. Haplotypes defined as neutral in this context were: (DR1/10)-DQB1*05:01, (DR8)-DQB1*04, (DR7)-DQA1*02:01-DQB1*02, (DR9)-DQA1*03-DQB1*03:03 and (DR13)-DQB1*06:03. DR molecules marked in parentheses are deduced based on the strong linkage disequilibrium. Longitudinal mother and offspring stool samples from five time points were analyzed including the meconium sample collected in the delivery hospital and another sample collected at the age of 3 in the offspring, a maternal sample collected at the beginning of the third trimester (gestational sample, another maternal sample collected at delivery (D), and a third maternal sample collected 3 months post-delivery (T). Paired serum and stool samples from mothers and offspring child were collected at the age of 3 months in the offspring. Selected characteristics of the study subjects are shown in Table S1.

### Analysis of molecular metabolites

Forty μl of serum sample was mixed with 90 μl of cold MeOH/H2O (1:1, v/v) containing the internal standard mixture (Valine-d8, Glutamic acid-d5, Succinic acid-d4, Heptadecanoic acid, Lactic acid-d3, Citric acid-d4. 3-Hydroxybutyric acid-d4, Arginine-d7, Tryptophan-d5, Glutamine-d5, 1-D4-CA,1-D4-CDCA,1-D4-CDCA,1-D4-GCA,1-D4-GCDCA,1-D4-GLCA,1-D4-GUDCA,1-D4-LCA,1-D4-TCA, 1-D4-UDCA, PFOA-13C4, PFNA-13C5, PFUdA-13C2,,PFHxS-18O2, PFOS-13C4) for protein precipitation. The tube was vortexed and ultra-sonicated for 3 min, followed by centrifugation (10000 rpm, 5 min). After centrifuging, 90 μl of the upper layer of the solution was transferred to the LC vial and evaporated under the nitrogen gas to the dryness. After drying, the sample was reconstituted into 60 μl of MeOH: H2O (70:30).

An Agilent 1290 Infinity LC system coupled with 6545 Q-TOF MS interfaced with a dual jet stream electrospray (dual ESI) ion source (Agilent Technologies, Santa Clara, CA, USA) was used for the analysis. Chromatographic separation was carried out using an Acquity UPLC BEH C18 column (100 mm × 2.1 mm i.d., 1.7 μm particle size), fitted with a C18 pre-column (Waters Corporation, Wexford, Ireland). Mobile phase A consisted of H_2_O: MeOH (v/v 70:30) and mobile phase B of MeOH with both phases containing 2mM ammonium acetate as an ionization agent. The flow rate was set at 0.4 mL·min^-1^ with the elution gradient as follows: 0-1.5 min, mobile phase B was increased from 5% to 30%; 1.5-4.5 min, mobile phase B increased to 70%; 4.5-7.5 min, mobile phase B increased to 100% and held for 5.5 min. A post-time of 5 min was used to regain the initial conditions for the next analysis. The total run time per sample was 18 min. The dual ESI ionization source was settings were as follows: capillary voltage was 4.5 kV, nozzle voltage 1500 V, N_2_ pressure in the nebulized was 21 psi and the N_2_ flow rate and temperature as sheath gas was 11 L·min^-1^ and 379 °C, respectively. In order to obtain accurate mass spectra in MS scan, the m/z range was set to 100-1700 in negative ion mode. MassHunter B.06.01 software (Agilent Technologies, Santa Clara, CA, USA) was used for all data acquisition. Data pre-processing was performed using MZmine 2.53 and identification was done using an in-house spectral library, based on m/z and retention time.

Quantification of BAs and PFAS were performed using a 7-point internal calibration curve. The identification was done with a custom data base, with identification levels 1 and 2 identification, based on Metabolomics Standards Initiative.

### PFAS analyses

Sample preparation and analysis for PFAS was carried out as described previously (Salihovićet al. 2020)In short, 450 μL acetonitrile with 1% formic acid, and internal standards were added to 150 μL serum and samples subsequently treated with Ostro Protein Precipitation & Phospholipid Removal 96-well plate (Waters Corporation, Milford, USA) Ostro sample preparation in a 96-well plate for protein precipitation and phospholipid removal. The analysis of PFAS was performed using automated column-switching ultra-sperformance liquid chromatography-tandem mass spectrometry (UPLC-MS/MS) (Waters, Milford, USA) using an ACQUITY C18 BEH 2.1×100mm×1.7μm column and a gradient with 30% methanol in 2mM NH4Ac water and 2mM NH4Ac in methanol with a flow rate of 0.3 mL/min. Quantitative analysis of the selected analytes was performed using the isotope dilution method; all standards (i.e., internal standards, recovery standards, and native calibration standards) were purchased from Wellington Laboratories (Guelph, Ontario, Canada). The method’s detection limits ranged between 0.02-0.19 ng/mL, depending on the analyte. NIST SRM 1957 reference serum as well as in-house pooled plasma samples were used in quality control, and the results agreed well with the reference values. The sum of total PFAS was calculated in ng/ml.

### Gut microbiome analysis by shotgun metagenomics

These methods is adapted versions of descriptions in the related work (Yassouret al. 2018).

#### Sample collection and DNA extraction

The stool samples were collected at home or in the delivery hospital. The samples collected at home were stored in the households’ freezers (−20°C) until the next visit to the study centre. The samples were then shipped on dry ice to the EDIA Core Laboratory in Helsinki, where the samples were stored at −80°C until shipping to the University of Tampere for DNA extraction. DNA extractions from stool were carried out using the vacuum protocol of PowerSoil DNA Isolation Kit.

#### Metagenome library construction

Metagenomic DNA samples were quantified by Quant-iT PicoGreen dsDNA Assay (Life Technologies) and normalized to a concentration of 50 pg μL−1. Illumina sequencing libraries were prepared from 100-250 pg DNA using the Nextera XT DNA Library Preparation kit (Illumina) according to the manufacturer’s recommended protocol, with reaction volumes scaled accordingly. Batches of 24, 48, or 96 libraries were pooled by transferring equal volumes of each library using a Labcyte Echo 550 liquid handler. Insert sizes and concentrations for each pooled library were determined using an Agilent Bioanalyzer DNA 1000 kit (Agilent Technologies).

### Metagenomic sequencing

Metagenomic libraries were sequenced on the Illumina HiSeq 2500 platform, targeting ∼2.5 Gb of sequence per sample with 101 bp paired-end reads (number of reads per sample is mentioned in Table S1). Reads were quality controlled by trimming low-quality bases, removing reads shorter than 60 nucleotides. Potential human contamination was identified and filtered using the KneadData Tool, v0.5.1 with the hg19 human reference genome. Quality controlled samples were profiled taxonomically using MetaPhlAn 2.0 (Segataet al. 2012; Truonget al. 2015).

### Statistical analysis

The linear discriminant analysis effect size (LEfSe) (Segataet al. 2011) was applied to identify differentially abundant taxa between the groups (high vs. low exposure). Wilcoxon rank-sum tests in LEfSe were used to identify significant differences. Spearman correlation coefficients were calculated using the Statistical Toolbox in MATLAB 2017b and p values <0.05 (two-tailed) were considered significant for the correlations. The individual Spearman correlation coefficients (R) were illustrated as a heat map using the “corrplot” package (version 0.84) for the R statistical programming language (https://www.r-project.org/). The subsequent visualization were done using ggplot2 R package.

## Data Availability

Metagenomic sequencing data for all subjects have been deposited to the SRA database under BioProject ID PRJNA475246.The metabolomics datasets and the clinical metadata generated in this study were submitted to MetaboLights (Haug et al., 2020), under accession number MTBLS875.

## Declaration of Competing Interest

The authors declare that they have no known competing financial interests or personal relationships that could have appeared to influence the work reported in this paper.

## Acknowledgments

We would like to thank Larson Hogstrom (Broad Institute, United States) for providing the taxonomic table from the EDIA metagenomics dataset. This study was supported by the Swedish Research Council (grant no. 2016-05176 to T.H and M.O), Formas (grant no. 2019-00869 to T.H and M.O), and the Novo Nordisk Foundation (Grant no. NNF20OC0063971 to T.H. and M.O.). The EDIA study was supported by the National Institute of Diabetes and Digestive and Kidney Diseases (NIDDK), National Institutes of Health (No. 1DP3DK094338-01 to M.K.), the Academy of Finland Centre of Excellence in Molecular Systems Immunology and Physiology Research 2012-17, No. 250114 to M.K. and M.O.). Further support was received by the Academy of Finland postdoctoral grant (No. 323171 to S.L.) and the Medical Research Funds, Tampere and Helsinki University Hospitals (to M.K.).

## Notes

### Competing Interest Statement

The authors have declared no competing interest.

### Clinical Trial

NCT01735123

### Author Declarations

The study followed the guidelines of the Declaration of Helsinki for research on human participants. The study was carried out according to good clinical practice. The study protocol was approved by the Joint Municipal Authority of the Pirkanmaa Hospital District, Finland (no. R11166). The parents gave their written informed consent, which was overseen by the Ethical Committee of the Joint Municipal Authority of the Pirkanmaa Hospital District. The study is registered under Clinicaltrials.gov Identifier NCT01735123.

